# Building knowledge of university campus population dynamics to enhance near-to-source sewage surveillance for SARS-CoV-2 detection

**DOI:** 10.1101/2021.08.03.21261377

**Authors:** Chris Sweetapple, Peter Melville-Shreeve, Albert S. Chen, Jasmine M. S. Grimsley, Joshua T. Bunce, William Gaze, Sean Fielding, Matthew J. Wade

## Abstract

Wastewater surveillance has been widely implemented for monitoring of SARS-CoV-2 during the global COVID-19 pandemic, and near-to-source monitoring is of particular interest for outbreak management in discrete populations. However, variation in population size poses a challenge to the triggering of public health interventions using wastewater SARS-CoV-2 concentrations. This is especially important for near-to-source sites that are subject to significant daily variability in upstream populations. Focusing on a university campus in England, this study investigates methods to account for variation in upstream populations at a site with highly transient footfall and provides a better understanding of the impact of variable populations on the SARS-CoV-2 trends provided by wastewater-based epidemiology. The potential for complementary data to help direct response activities within the near-to-source population is also explored, and potential concerns arising due to the presence of heavily diluted samples during wet weather are addressed. Using wastewater biomarkers, it is demonstrated that population normalisation can reveal significant differences between days where SARS-CoV-2 concentrations are very similar. Confidence in the trends identified is strongest when samples are collected during dry weather periods; however, wet weather samples can still provide valuable information. It is also shown that building-level occupancy estimates based on complementary data aid identification of potential sources of SARS-CoV-2 and can enable targeted actions to be taken to identify and manage potential sources of pathogen transmission in localised communities.

## 1. INTRODUCTION

Wastewater-based epidemiology (WBE) is a promising tool for complementary surveillance of infectious diseases and provision of early warning of disease outbreaks (Sims and Kasprzyk-Hordern 2020), and has received considerable interest during the global COVID-19 pandemic (e.g. Bivins et al. 2020, Gonzalez et al. 2020, Polo et al. 2020). Since a significant proportion of individuals infected with the severe acute respiratory syndrome coronavirus 2 (SARS-CoV-2) shed the virus’ ribonucleic acid (RNA) in their faeces (Medema et al. 2020), SARS-CoV-2 RNA concentrations in wastewater can be used to provide an indicator of the disease prevalence without relying on clinical testing data. Wastewater networks can, thus, be viewed as a reflection of the microbiome of the population using the upstream systems (Newton et al. 2015). This is beneficial as wastewater surveillance is independent from people participating in testing (a testing bias towards symptomatic individuals and testing reluctance may result in underrepresentation of the infected in clinical test data).

To date, SARS-CoV-2 related wastewater surveillance projects have been implemented in at least 54 countries, covering more than 2,000 different sites (University of California 2021). The European Union has recognised WBE as a tool to address emerging and future public health issues (European Commission 2020), and Member States have been mandated to engage with the development of the European Sewage Sentinel System for SARS-CoV-2, which will provide systematic surveillance of SARS-CoV-2 and its variants in EU wastewaters (Gawlik et al. 2021). In England, WBE for SARS-CoV-2 surveillance, led by the Joint Biosecurity Centre and Defra Group under the Environmental Monitoring for Health Protection (EMHP) programme, covers in excess of 500 sites as of June 2021.

Wastewater samples are commonly collected at sewage treatment works (STWs), thereby providing an indicator of SARS-CoV-2 prevalence across the entire STW catchment – in the Netherlands, for example, samples are taken daily at every STW in the country and analysed for presence of SARS-CoV-2 (Dutch Water Sector, 2020). However, in-network or near-to-source sampling can provide greater resolution and, potentially, additional insights: If the population upstream of the sampling point is smaller, then better targeted actions can be taken to address and mitigate any outbreak detected by monitoring of the wastewater. Application of wastewater surveillance at a building scale, for example, can aid management of outbreaks in discrete populations, and may be beneficial in high risk settings such as schools, prisons and critical points in the food supply chain (Wade et al. 2020).

Near-to-source wastewater monitoring, employed at a hospital building, was able to detect a single asymptomatic individual among as many as 400 residents (Karthikeyan et al. 2021). The same study also found that trends in the number of cases could be successfully captured, with a strong correlation between the wastewater SARS-CoV-2 concentration (gene copies (gc)/l) and the number of active COVID-19 patients identified.

However, challenges remain in the interpretation and use of wastewater SARS-CoV-2 data as an indicator of prevalence – one of which is the impact of variation in the upstream population size. The importance of accounting for fluctuating population sizes in WBE has previously been highlighted and investigated in the context of applications such as illicit drug monitoring (Been et al. 2014). However, in wastewater SARS-CoV-2 monitoring, the value considered as an indicator of prevalence is still typically reported as a concentration (gc/l) (i.e. not normalised with respect to population like prevalence values) (e.g. Karthikeyan et al. 2021, Saththasivam et al. 2021, Prado et al. 2021), or even just a binary presence or absence of SARS-CoV-2 RNA (e.g. Gibas et al. 2021). Using concentration as an indicator of prevalence may be reasonable for monitoring efforts at a city-scale (i.e. those at a STW) when the population size and wastewater dilution are relatively static, as a constant average SARS-CoV-2 load per capita would then correspond to a constant SARS-CoV-2 concentration (assuming constant wastewater production per capita). It is not valid, however, when population is highly variable and/or there are significant dilution events, since a constant average SARS-CoV-2 load per capita would then yield a variable SARS-CoV-2 concentration. This is illustrated in Figure 1, which shows the theoretical relationship between SARS-CoV-2 concentration, SARS-CoV-2 load per capita, population size and baseflow (i.e. flow not attributed directly to the population), based on a mass balance approach and assuming a per capita wastewater production of 150 l/d (further details in the Supplementary Information).

**Figure 1.**
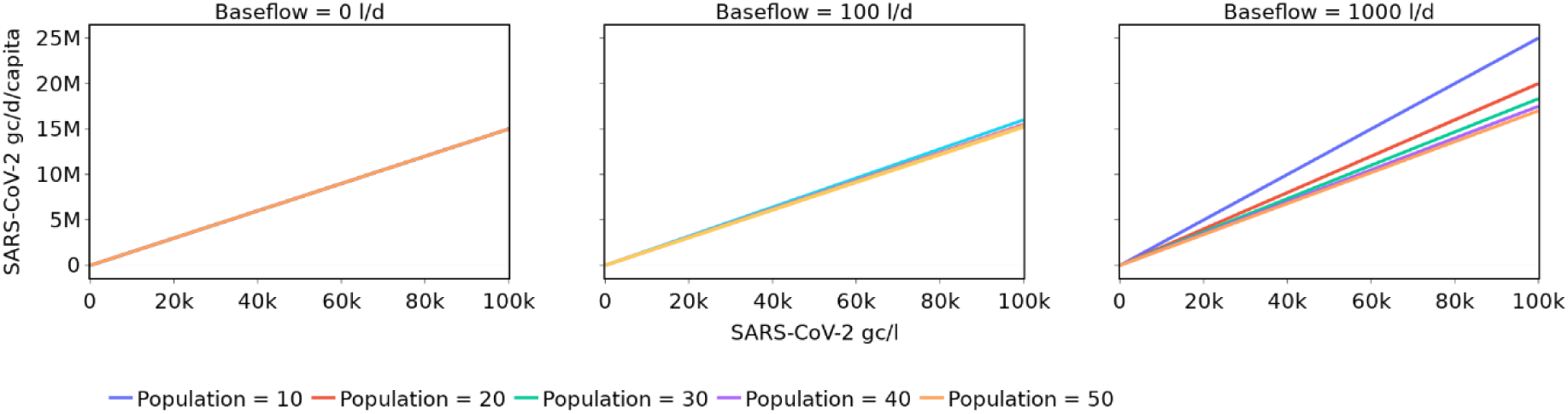
Relationship between SARS-CoV-2 concentration and daily load per capita for different populations and base flows

The impacts of variable population are likely to be exacerbated when interpreting the results for near-to-source sites, since population movement will not be averaged out to the same extent that it is at the STW level. This is especially important for near-to-source sites that are subject to significant daily variability in upstream population (for example due to a lack of activity at weekends or outside of academic term time at educational sites), and approaches are required to ensure that the effects of variable populations are not overlooked.

The first aim of this study, therefore, is to investigate methods by which the effects of variation in the upstream population at a highly dynamic near-to-source site can be accounted for, and to build a better understanding of the impact of highly variable populations at a near-to-source site on the insights into SARS-CoV-2 prevalence from WBE. Ammoniacal nitrogen (NH_3_-N) and orthophosphate (PO_4_^3-^) in the wastewater are considered as potential indicators of population dynamics and for population normalisation. The work presented here is specifically focused on a university campus case study, where footfall varies considerably throughout the year (especially during periods of lockdown), and where knowledge of COVID-19 prevalence provided by WBE has significant potential to target actions to identify positive cases and reduce transmission between the university and the wider community.

Whilst near-to-source surveillance captures wastewater from a smaller and better-defined population than monitoring at a STW level, the pool of potential candidates for the source of any SARS-CoV-2 detected may still be large. The university monitored in this study, for example, currently has approximately 20,900 students based at the campus and over 4,300 staff members. As such, the insights provided by WBE would be of greater assistance to campus managers if the potential source(s) could be narrowed down further, e.g. to the most probable building or buildings on site, so that better targeted public health actions may be taken. The second aim of this study, therefore, is to investigate the potential of complementary data to help direct response activities to the most appropriate locations within the site from which wastewater is collected. This is achieved with the use of toilet flush data, collated at washroom and building levels, which provide information on the major sources of wastewater at the time of SARS-CoV-2 detection and the relative activity levels at different locations on the campus.

Lastly, this study identifies samples collected during wet and dry weather periods to enable exploration of the impact of wet weather periods on wastewater SARS-CoV-2 concentrations and population normalisation. This aims to establish the value of wastewater monitoring in wet versus dry periods, and address potential concerns arising due to the presence of heavily diluted samples during wet weather.

It is intended that the results of this study will give improved confidence in the SARS-CoV-2 trends observed in wastewater from near-to-source sampling sites, and will enable better targeted actions to identify and manage potential sources of pathogenic transmission in localised communities.

## 2. METHODS

### 2.1. Case study site selection

The University of Exeter’s Streatham Campus was monitored as a near-to-source pilot as part of the UK National Surveillance Programme. Wastewater samples were collected in three locations at the university: Two were downstream of student residential accommodation, and one downstream of the main campus (consisting of multiple academic, administrative and social buildings). This study focuses on the main campus monitoring site. The results are expected to be of particular benefit here, since the number of people on campus is much more variable than in accommodation blocks (due to working patterns and lecture scheduling, for example), and population normalisation will thus have greater impact on the understanding of prevalence. The maximum size of the potential upstream population is also considerably larger, given that a large proportion of the university’s 25,000 students and staff will have access to the main campus buildings (although not at the same time), whereas only a small proportion will be associated with individual accommodation sites. As such, any insights provided by this study that enable the potential source of SARS-CoV-2 detected in the wastewater to be narrowed down further will be much more valuable for the larger, main campus site.

Furthermore, whilst the university does hold data on the number of students and staff that have tested positive for SARS-CoV-2, there are no data on the total number of students and staff using the campus each day (or whether those that have tested positive have actually been on the main campus site on a given day) and, thus, prevalence cannot be calculated from existing sources of information.

### 2.2. SARS-CoV-2 population normalisation

The daily wastewater SARS-CoV-2 load per capita (*L*_*d*_, gc/capita/day) (i.e. a value that is comparable with prevalence) can be calculated using Eq. 1, where *C*_*d*_ is the SARS-CoV-2 concentration (gc/l), *Q*_*d*_ is the daily flow rate (l/day), and *N*_*d*_ is the population size (in this case, the number of individuals using the university campus).

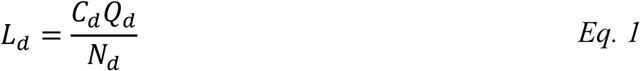

However, the population size on any given day is unknown and must, therefore, be estimated if this equation is to be used.

There are multiple potential approaches to population estimation. In WBE, biomarkers (substances excreted by humans) that have homogeneous excretion throughout a community at low variance can be used as indicators of population size (Choi et al. 2018). These include, for example, creatinine and coprostanol (Daughton 2012), and the cross-assembly phage (crAssphage) (Wu et al. 2021). Water quality parameters, such as biochemical oxygen demand (BOD), chemical oxygen demand (COD), total phosphorus (TP), total nitrogen (TN) and ammonia or ammonium, may also be considered as population biomarkers (Xagoraraki and O’Brien 2019, Nuijs et al. 2011): Been et al. (2014), for example, used measured ammonium concentrations in conjunction with the expected daily per capita ammonium discharge to estimate population size for the purposes of illicit drug monitoring, and Rico et al. (2017) generated population estimates based on TN, TP, BOD and COD and typical daily per capita discharges for each parameter.

Provided that daily flow rate (*Q*_*d*_, l/day) and biomarker concentration (*X*_*d*_, mg/l) data is available for a historical period with known population (*N*_*d*_), the daily discharge per capita of a biomarker (*x*, mg/capita/day) can be estimated using Eq. 2, and the population on any given day using Eq. 3.

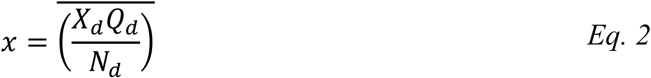

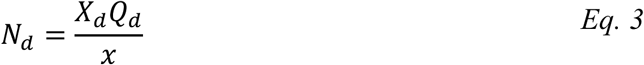

Once the daily discharge per capita of a specific biomarker is known, concentrations of a substance of interest (in this case, SARS-CoV-2 RNA) can then be population normalised using Eq. 4, without calculating the population size and without any ongoing requirement for flow rate data (based on substitution of Eq. 3 into Eq. 1):

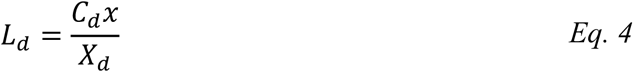

Where *L*_*d*_ is the daily per capita load of SARS-CoV-2 at day *d* (gc/capita/day) and *C*_*d*_ is the SARS-CoV-2 concentration in the wastewater at day *d* (gc/l).

Population estimates may also be generated using non-wastewater data: Thomas et al. (2017) and Deville et al. (2014), for example, produced dynamic population estimates using mobile phone communication data. With respect to the case study site in particular, there is ongoing investigation into the potential use of wi-fi tracking for population monitoring under the ‘*Riba to Reality*’ project (UKRI 2020); whilst there are no results available for use from this yet, it is a promising development. Additional data available for the case study site that may be indicative of population includes metered water and electricity supply and washroom-level flush counts.

Each of these methodologies is subject to limitations, however, and there are some barriers to implementation in a near-to-source study, where it should be noted that not everyone on site will contribute to the wastewater collected since some visits may only be short. Whilst mobile phone-based population estimates have the advantage that they can account for people who spend only a short period of time in the monitored location, irrespective of whether they produce any wastewater, this may be a disadvantage in near-to-source WBE as the population estimate is likely to exceed the number of people that contribute to the wastewater being sampled, and thus result in an underestimate of SARS-CoV-2 gene copies per capita. To avoid this problem, a population normalisation methodology based on wastewater characteristics is considered more appropriate for a near-to-source site with a highly dynamic population.

For other non-wastewater indicators such as metered water, electricity supply and flush counts, data is only available for a subset of buildings on campus. As there is insufficient evidence that the occupancy dynamics of these buildings are representative of those of the whole campus, it is inappropriate to use these for population normalisation (although building-level population estimates may aid SARS-CoV-2 source identification, as discussed in Section 2.3).

There are also potential disadvantages of using biomarkers in wastewater for population normalisation. For the case study site, ammoniacal nitrogen and orthophosphate concentrations are available, but it is recognised that nutrient concentrations will be affected by industry (Xagoraraki and O’Brien, 2019) and, where there is significant industrial input into the sewer network, this may lead to errors in population estimates (Nuijs et al. 2011). Other studies, however, have concluded that use of such nutrients is appropriate (e.g. Choi et al. 2018, Been et al. 2014), and Zheng et al. (2017) found population estimates based on ammonia-nitrogen to show good agreement with estimates provided by wastewater treatment plant operators with local knowledge. Furthermore, given that the focus of this study is a near-to-source site, and it is known with a high degree of certainty that there are no industrial inputs to the wastewater, these concerns are not considered a barrier in this study.

A remaining issue with this approach is the need to calculate the daily biomarker load excreted per capita. Whilst previous studies have either estimated a site-specific value using historical data (e.g. Rico et al. 2017) or used published values from the literature (e.g. Nuijs et al. 2011), neither of these approaches is appropriate for this study. To calculate a site-specific value, data is required from a period with known population; however, no wastewater data was collected at the case study site prior to the pandemic (when the campus could be assumed to be at full capacity, i.e. a known population size), and occupancy figures during the period of wastewater monitoring are unknown. Use of daily values from literature is also inappropriate since these are typically calculated at a STW level and thus capture discharge from every occupant over a full 24 hours, whereas campus users are only present for part of the day and their biomarker discharge will, therefore, be proportionately smaller.

Based on Eq. 4, and given that the daily biomarker discharge per capita (*x*) is constant at a given site but cannot be calculated, this study calculates values that are proportional to the SARS-CoV-2 load per capita (gene copies per mg of biomarker) instead of the actual load per capita (gene copies per capita) (Eq. 5). These values are not comparable between sites; however, they are sufficient for understanding the effects of population normalisation on SARS-CoV-2 trends at a single site. Ammoniacal nitrogen and orthophosphate are both considered as potential biomarkers.

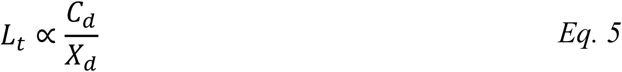

### 2.3. Source identification

As discussed in Section 2.2, supplementary data, including metered water supply, electricity supply and flush counts, are available for a subset of buildings on campus, and may be used to analyse occupancy dynamics within these buildings. Preliminary investigation shows very poor agreement between building-level population estimates based on metered water and electricity supply at the case study site (see Supplementary Information for example) – potentially due to seasonal and other non-population-related influences – and, therefore, only flush counts are considered further.

Given that flush count data relates directly to the production of wastewater, this information can be used to assess the relative contribution of different buildings to the wastewater sampled, and thus provide additional insights into the potential source(s) of any SARS-CoV-2 RNA detected in the wastewater.

It has previously been shown that the number of flushes per occupant varies by building (Melville-Shreeve et al. 2021); therefore, flush counts cannot be compared directly between buildings and instead are used to estimate the occupancy of each building on any given day. The maximum capacity of each building in which flush counts were monitored is shown in Figure 2, and it is assumed that on weekdays during the autumn term of 2019 (pre-pandemic) all buildings were operating at full capacity. The mean daily flush count per capita is then estimated for each building using Eq. 6, and building occupancies during the wastewater monitoring period are estimated using Eq. 7.

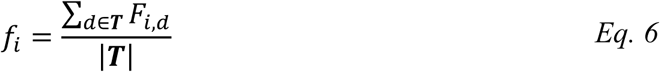

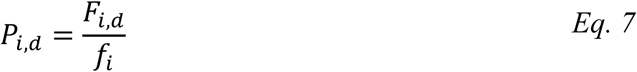

**Figure 2.**
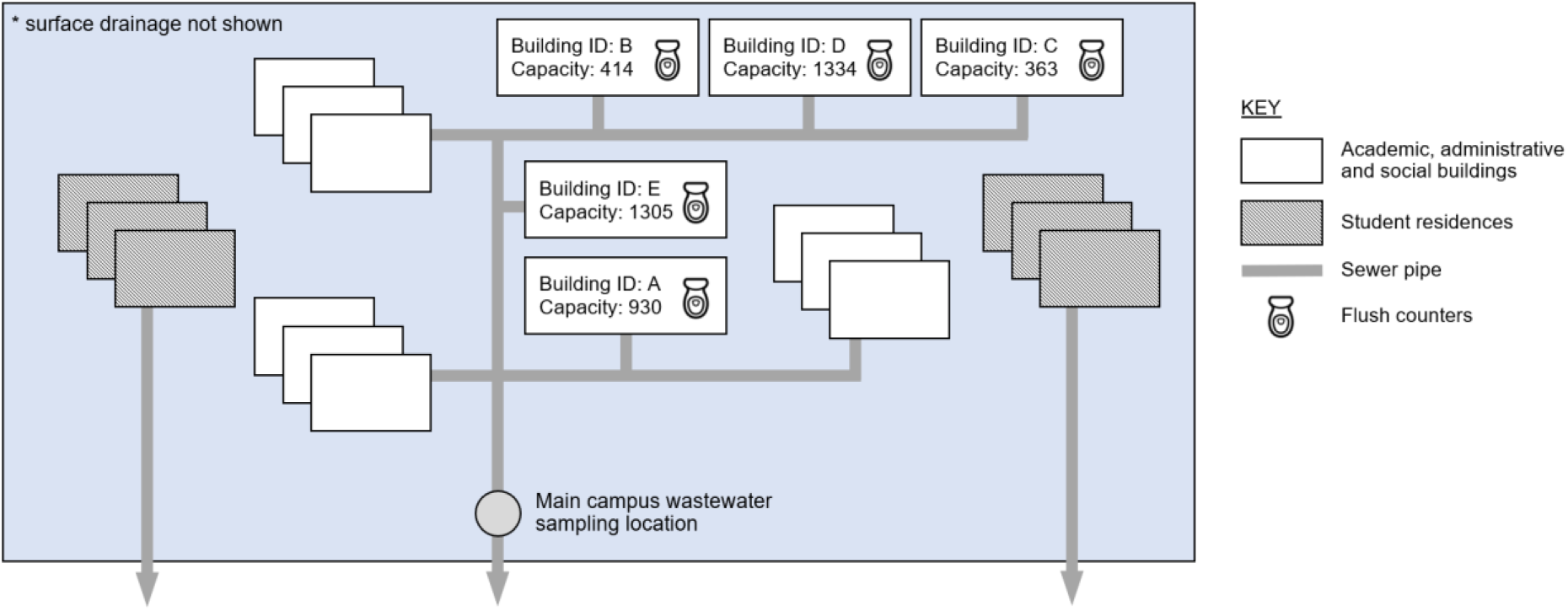
Case study site schematic, illustrating sampling locations and wastewater flow from campus buildings. Surface drainage is omitted for clarity.

Where *f*_*i*_ is the mean daily per capita flush count for building *i, F*_*i,d*_ is the total flush count for building *i* on day *d*, ***T*** denotes the set of term time days where building occupancy is assumed to equal building capacity, and *P*_*i,d*_ is the estimated population of building *i* on day *d*.

This methodology is not used to estimate the total population of the campus on a daily basis for the purposes of population normalisation as only as subset of the buildings have flush counts monitored. As significant variability is found between buildings for the daily per capita flush counts and the population estimates (see Section 3.3), it is not considered appropriate to assume that the population dynamics for the monitored buildings are indicative of those for the whole campus. Based on flush count data, it is not possible to either eliminate or target any of the buildings that are not monitored as a potential source of any SARS-CoV-2 detected in the wastewater; however, the population estimates provided can be used to prioritise/deprioritise each of the buildings with flush count data available in the search for the source of the SARS-CoV-2. A set of flush data for every toilet on campus would yield additional value as a future extension to this study.

### 2.4. Wet and dry weather days

Days are classified as wet weather or dry weather (or neither) based on Environment Agency rainfall data for the nearest rain gauge (station ID 45184, located approximately 4km from the case study site) (Environment Agency, 2021). A day is considered a dry weather day if no rainfall has been recorded on that day or in the previous six hours. A day is considered a wet weather day if the total rainfall for that day exceeds 2mm (with the threshold set based on the depth of rainfall expected to cause runoff from impervious surfaces (Ladson 2019)). Days that fall into neither category (i.e. have rainfall but less than 2mm, or have had rainfall in the preceding six hours) are not classified as either wet or dry.

### 2.5. Data collection and handling

This study incorporates both wastewater data and supplementary data relating to occupancy of the campus, and a schematic of the case study site illustrating the data collection locations is given in Figure 2. Wastewater samples were collected downstream of the academic, administrative and social buildings (wastewater from student residences on campus are not contributory), and they were analysed to provide concentrations of SARS-CoV-2, ammoniacal nitrogen and orthophosphate. Wastewater flow rate was measured at the same location. Flush counts were collected from five buildings, which have a combined occupancy of 4,346 at full capacity.

Further detail on the data collection methodologies and data handling for each data type is provided in the following sections.

#### 2.5.1. Wastewater characteristics

Methodologies for the SARS-CoV-2, ammoniacal nitrogen and orthophosphate data collection and processing are summarised below. Further details are available in Wade et al. (2020) and Hoffman et al. (2021).

Composite wastewater samples were collected over a 24-hour period using HACH AS950 autosamplers (100ml every 15 minutes); these were then retrieved and kept at 4°C to prevent degradation of RNA during transportation to a laboratory for analysis. Quantitative Polymerase Chain Reaction with a reverse transcriptase step (RT-qPCR) was used to quantify the N1 gene from the SARS-CoV-2 virus in the wastewater samples. This provides a measurement of the number of RNA copies in the sample, which is reported as gene copies per litre of wastewater sample collected. The practical limit of detection (LOD) is 133 gc/l. No adjustments for analytical efficiency are applied.

Concentrations of ammoniacal nitrogen and orthophosphate were determined using colorimetric assays. In both cases, outliers (more than three standard deviations from the mean) are omitted from further analyses, since these may originate from a sample that was poorly mixed and thus unrepresentative of the average wastewater composition at that time. These represent 0% of the ammoniacal nitrogen measurements and 1% of the orthophosphate measurements (two samples) in the analysis period.

The level and velocity of wastewater in the sewer were monitored at two-minute intervals in the same location as collection of wastewater samples, and were used to calculate wastewater flow rates. To enable comparison with the wastewater constituent concentrations measured in the composite samples, flow data is resampled to provide daily values.

#### 2.5.2. Flush counts

Flush monitoring systems were installed in washrooms at the University of Exeter, as described by Melville-Shreeve et al. (2021a, 2021b). These captured flush data for washrooms in six buildings, covering a total of 38 washrooms, and provided real time flush counts for 119 toilets (approximately 18% of toilets on campus). Wheelchair-accessible washrooms were omitted for operational reasons, but all other washrooms in the selected buildings were monitored. The selected buildings represent typical university department buildings. The monitoring system provided high resolution data, with either a ‘zero’ (no flush) or a ‘one’ (flush) recorded every minute for every toilet.

In the case of any faults in the system, gaps may appear in the data and give the impression of a reduced daily flush count if not identified and accounted for. Therefore, for quality assurance purposes, daily data completeness (the total number of data points collected, expressed as a percentage of the expected number of data points for the time period) is evaluated at a washroom level. Since estimating flush counts for periods with insufficient completeness would provide results with unknown accuracy, all washrooms with less than 90% data completeness during the wastewater monitoring period are omitted from further analysis. This leaves 22 washrooms, covering five buildings, as illustrated in Figure 2. Additionally, any day with less than 90% data completeness in any washroom is omitted from further analysis. No outliers (more than three standard deviations from the mean) are present in the total daily flush counts.

Detailed data completeness results for individual washrooms are provided in the Supplementary Information (Figure S1), and days omitted from analyses due to insufficient site-level flush count data availability are identified in Figure 3, Section 2.6.

**Figure 3.**
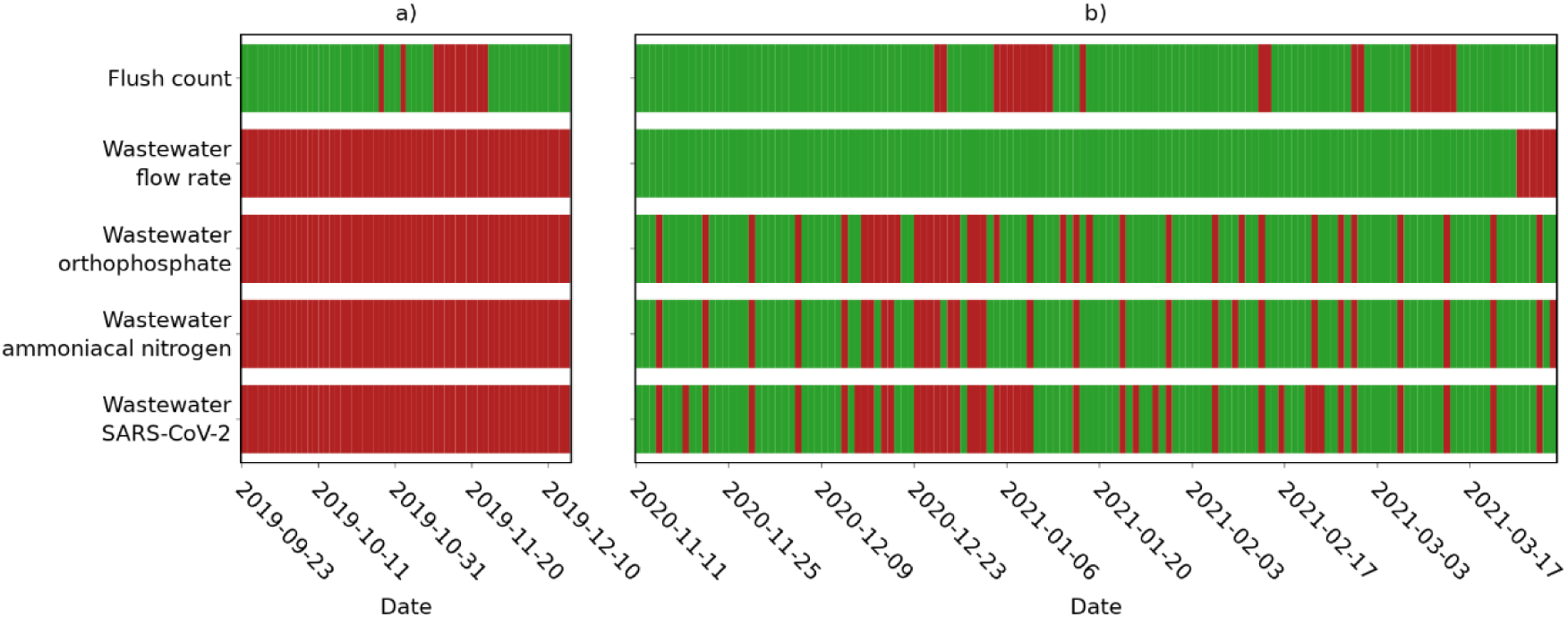
Daily data availability after cleaning, a) during a period of expected full occupancy, and b) for the wastewater analysis period. Red indicates periods of missing data.

Since flush counts are required to provide an indication of building occupancy, flushes attributable to cleaning are identified and removed from the daily building level totals (as these are not related to the building occupancy and do not contribute to SARs-CoV-2, ammoniacal nitrogen or orthophosphate measured in the wastewater). A washroom is assumed to have been cleaned if every toilet in the washroom has been flushed in a 15 minute period at least once during the day; the number of flushes attributed to cleaning (a maximum of one per toilet per day) is then subtracted from the total flush count. Further detail on the impact of flush count adjustments for cleaning is provided in the Supplementary Information.

### 2.6. Data availability and key dates

Wastewater SARS-CoV-2, ammoniacal nitrogen and orthophosphate data is available from 11^th^ November 2020, and sampling is ongoing. Calibrated wastewater flow rate data is available from the same date, up until 23^rd^ March 2021. Within these periods, wastewater constituent data is typically available for six days a week, although sampling was reduced during the university’s Christmas break (12^th^ December 2020 – 3^rd^ January 2021), and flow rate data is available for every day. Flush data was collected from 1^st^ July 2019 until 29^th^ March 2021, although gaps are present where days have been omitted due to insufficient data completeness.

The period 11^th^ November 2020 to 29^th^ March 2021, i.e. when both wastewater constituent and flush data is available, is selected for analysis of the impact of population dynamics and SARS-CoV-2 trends. Additional flush data from a period with assumed full occupancy (weekdays during the 2019 autumn term) is also used in the source identification, as detailed in Section 2.3.

Figure 3 shows the availability of each data source over these two periods, with data gaps shown in red. Key dates within the wastewater monitoring period which may affect the population on campus are summarised in Table 1.

**Table 1.** Key dates in the wastewater SARS-CoV-2 analysis period

| Date | Description | Abbreviation |
| --- | --- | --- |
| 2<sup>nd</sup> December 2020 | End of second national lockdown | NL2-E |
| 11<sup>th</sup> December 2020 | End of university term | T-E |
| 24<sup>th</sup> December 2020 | Start of university closure period | CP-S |
| 1<sup>st</sup> January 2021 | End of university closure period | CP-E |
| 4<sup>th</sup> January 2021 | Start of university term | T-S |
| 6<sup>th</sup> January 2021 | Start of third national lockdown | NL3-S |

## 3. RESULTS AND DISCUSSION

### 3.1. Population dynamics

Population dynamics indicated by ammoniacal nitrogen and orthophosphate are analysed to check that they appear plausible and to confirm that these biomarkers are a reasonable basis for population normalisation for the case study site.

Site-specific values for the daily discharge per capita of ammoniacal nitrogen and orthophosphate are unknown and cannot be calculated with the available information, and thus absolute population cannot be estimated. The total biomarker load is, however, proportional to population (based on Eq. 3) and can be used to illustrate population dynamics and trends. Daily loads of ammoniacal nitrogen and orthophosphate, each of which is expected to be proportional to the population on campus, are therefore shown in Figure 4. These are calculated using the measured wastewater flow rate and biomarker concentration (*Q*_*d*_*X*_*d*_).

**Figure 4.**
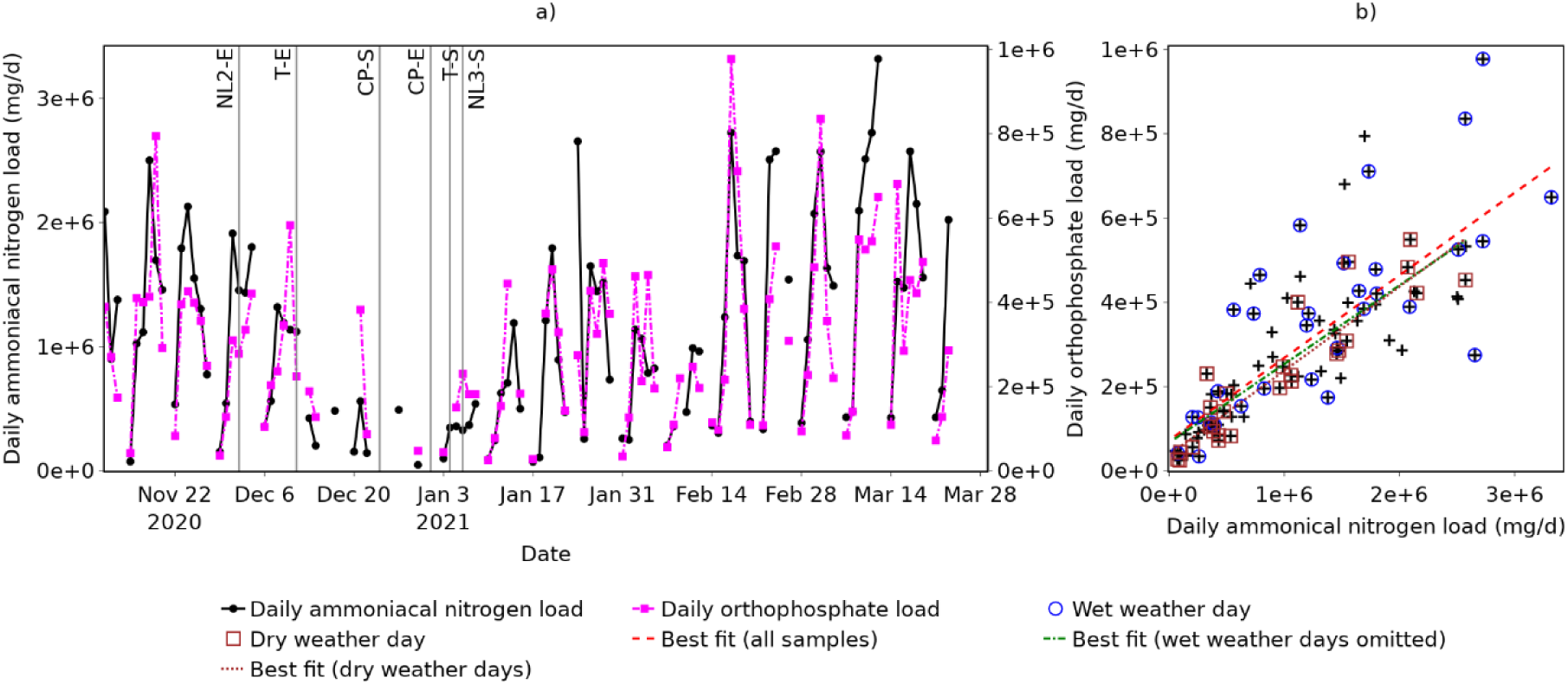
Daily ammoniacal nitrogen and orthophosphate loads, showing: a) change over time; and b) correlation. Key dates are indicated using abbreviations defined in Table 1.

Figure 4a suggests that population is highly variable, whichever biomarker is considered. Two general patterns are observed: Firstly, a seven-day cycle, with daily biomarker loads typically low (<500,000 mg ammoniacal nitrogen or <200,000 mg orthophosphate) at weekends and higher during the week; and secondly, a longer term trend with levels dropping throughout December and rising through January. This matches what would be expected, since the main campus was not used for lectures at weekends and fewer staff would have been working. In December, the population on campus would be expected to drop towards, and following, the end of term (T-E in Figure 1) as students started to return home and lectures ceased. Few samples were taken during the Christmas closure period (CP-S to CP-E), when the population on campus would have been close to zero, but where available the biomarker loads are typically representative of a low population. Population would then be expected to rise following the end of the Christmas closure period (CP-E) and start of term (T-S), although remain somewhat suppressed due to the start of the third national lockdown (NL3-S), and this is reflected in an increase in the weekday biomarker loads.

The correlation between ammoniacal nitrogen and orthophosphate loads (Figure 4b) is strong but not perfect (Pearson correlation coefficient *r* = 0.792), indicating that the population estimates and population normalisation results yielded by each will differ. This may be attributed to variations in the per capita discharge of each biomarker and the potential presence of additional sources of either, or both, and is not unexpected given previous studies have also reported discrepancies between population estimates based on different wastewater constituents (e.g. Nuijs et al. 2011). Given that no population estimates from alternative sources are available for reference, it is not possible to determine whether ammoniacal nitrogen or orthophosphate provides a more accurate representation of population dynamics and, therefore, both are considered in the following normalisation (Section 3.2).

If samples taken during periods of wet weather are omitted, the correlation increases marginally (*r* = 0.804), and if only samples taken on dry weather days are included, the correlation increases further (*r* = 0.917). This suggests that confidence in population estimates and population normalisation should be greatest during dry weather days; however, population normalisation is not restricted to these days since this would prevent normalisation of over 70% of samples, and there is still reasonable agreement between population estimates based on the different biomarker loads even on wet weather days (*r* = 0.758).

Due to the topology and geology of the site (steep sandstone and mudstone), ingress to the sewer (infiltration of groundwater) is not expected to be a major contributor to the wastewater sampled; however, a similar study in an area with a high water table and a poorly maintained pipe network may yield different results due to the increased impact of wet weather periods.

### 3.2. Impact of population normalisation on wastewater SARS-CoV-2 trends

SARS-CoV-2 RNA concentrations measured in wastewater samples at the case study site over the period 11^th^ November 2021 to 29^th^ March 2021 are shown in Figure 5 (blue lines). Samples with a concentration below the LOD are displayed with a value of 20 gc/l so that general trends can be observed; this value is half the LOD and selected based on the assumption that all values below the LOD are equally probable. The concentrations in Figure 5 show that there is only intermittent detection of SARS-CoV-2 during the monitoring period (13% of samples collected contain a detectable level of SARS-CoV-2), with the most frequent detection and the maximum concentration both occurring in December 2020.

**Figure 5.**
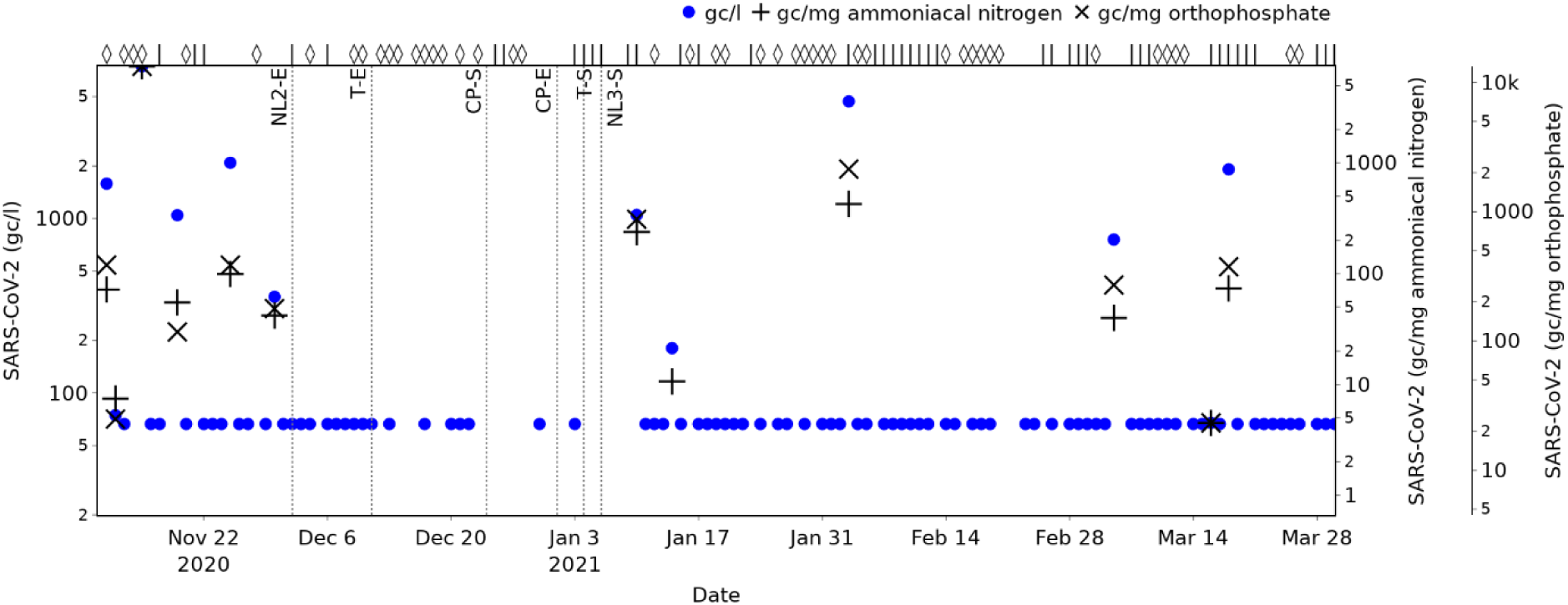
Wastewater SARS-CoV-2 concentrations and normalised values over the wastewater monitoring period. Wet weather days are indicated with a ‘⋄’ symbol, and dry weather days with a ‘|’ (top axis).

All SARS-CoV-2 RNA concentrations above the LOD are normalised with respect to ammoniacal nitrogen and orthophosphate, and shown with ‘+’ and ‘×’ symbols respectively in Figure 5, providing values that are expected to be directly proportional to the daily SARS-CoV-2 load per capita. To aid comparison of relative magnitudes with different units, y-axes are scaled so that the maximum gc/l, gc/mg ammoniacal nitrogen and gc/mg orthophosphate (all on 15^th^ November 2020) all appear at the same level, and similar for the minimum detected values (all on 16^th^ March 2021).

Figure 5 shows that population normalisation (whether based on ammoniacal nitrogen or orthophosphate) changes the picture of SARS-CoV-2 prevalence trends provided by the wastewater monitoring. The wastewater samples taken on 19^th^ November 2020 and 10^th^ January 2021, for example, have very similar SARS-CoV-2 RNA concentrations (1045 and 1048 gc/l respectively), but there is significant difference in their normalised values: Based on these, the daily per capita load is shown to be 333 % (based on ammoniacal nitrogen for normalisation) or 644% (based on orthophosphate for normalisation) higher on 10^th^ January than on 19^th^ November. This compares with an increase of just 0.3% in concentration. Neither of these samples were taken on a wet weather day, so the change cannot be attributed to dilution effects. This suggests, therefore, that population normalisation can have significant impact on the understanding of the relative severity of peaks in SARS-CoV-2, and that the presence of similar SARS-CoV-2 RNA concentrations may give a false impression of similar levels of prevalence.

This is supported by analysis of the rankings of the SARS-CoV-2 RNA concentrations and population normalised values. Whilst the rankings based on gc/mg ammoniacal nitrogen and gc/mg orthophosphate both have a strong correlation with the SARS-CoV-2 RNA concentration (Spearman’s rank correlation coefficients of 0.95 and 0.91 respectively), the sample on 10^th^ January is an example of where population normalisation significantly changes the ranking (from 6^th^ / 12 based on concentration, to 3^rd^ based on gc/mg ammoniacal nitrogen or gc/mg orthophosphate) and may alter the importance placed on the measurement and any response actions that may be considered.

A comparison of concentrations and population normalised values for all samples with a detectable level of SARS-CoV-2 is presented in Figure 6, illustrating more clearly the relationship between each metric and which measurements correspond to dry and wet weather days. In each case, linear lines of best fit and Pearson correlation coefficients shown are calculated based on the log_10_ values.

**Figure 6.**
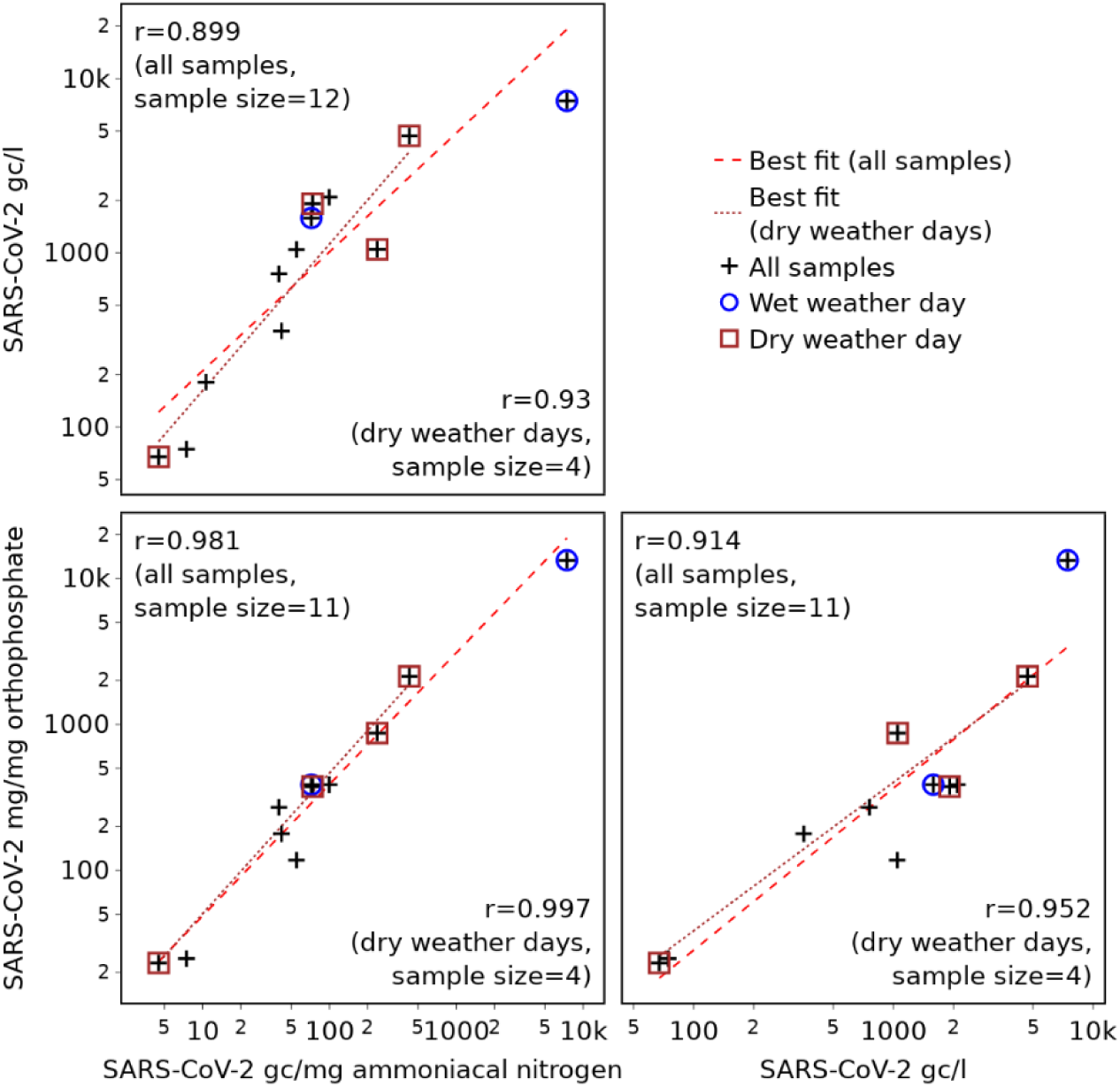
Correlations between wastewater SARS-CoV-2 concentrations and normalised values.

Figure 6 shows that, although strongly correlated, the SARS-CoV-2 concentration (gc/l) is perfectly linearly related to neither the gc/mg ammoniacal nitrogen (r = 0.899) nor the gc/mg orthophosphate (r = 0.914). This reinforces the assertion that population normalisation alters the SARS-CoV-2 prevalence trends provided by WBE, since the relative magnitude (with respect to the full data set) of all SARS-CoV-2 concentrations and normalised loads would be unaltered only if there is a perfect, linear relationship (r = 1) between SARS-CoV-2 concentration and the load per unit mass of biomarker. Whilst it may appear that the difference in the trends is likely to be minor, due to the strong correlation, the difference between the linear best fit lines and the normalised values is in fact considerable (Root Mean Square Percentage Error (RMPSE) = 83.7% for SARS-CoV-2 gc/mg ammoniacal nitrogen, and RMSPE = 89.7% for SARS-CoV-2 gc/mg orthophosphate.

Comparison of the two population normalised values (gc/mg ammoniacal nitrogen and gc/mg orthophosphate) shows that these exhibit a very strong correlation (r = 0.981), suggesting that both will provide a similar understanding of trends in daily per capita loads of SARS-CoV-2.

Given that there is greater confidence in population normalisation using samples from dry weather days (Section 3.1), correlation coefficients are also calculated based on dry weather samples only. Again, these show there to be a very strong (nearly perfect) correlation between the two population normalised metrics (r = 0.997), and a weaker correlation between these and the SARS-CoV-2 RNA concentration (r = 0.930 and 0.952 for normalisation based on ammoniacal nitrogen and orthophosphate respectively). These correlation coefficients are based on only four samples (as this is the total number of samples with a SARS-CoV-2 concentration above the LOD collected on dry weather days) and, therefore, the confidence intervals (detailed fully in the Supplementary Information, Table S1) are wider than for the correlation coefficients based on all samples. However, the correlation coefficients still support the conclusion that population normalisation alters the SARS-CoV-2 trends provided by WBE, as there is not a perfect linear relationship between SARS-CoV-2 concentration and SARS-CoV-2 normalised by either biomarker. They also support the suggestion that normalisation using either metric is similarly beneficial, as the correlation between SARS-CoV-2 gc/mg ammoniacal nitrogen and gc/mg orthophosphate is very strong even when considering the full confidence interval (0.867 ≤ r ≤ 1.000 for a significance level of 0.05).

### 3.3. Potential sources of SARS-CoV-2

Estimated daily flush counts per occupant for each of the monitored buildings during the period of assumed full occupancy, as calculated using Eq. 6 and required to estimate dynamic building occupancies for SARS-CoV-2 source identification, are summarised in Table 2. The total flush counts during the assumed full occupancy period, from which these values are calculated, are provided in the Supplementary Information (Figure S2). The coefficients of variation indicate that there will be a high degree of certainty in population estimates based on the estimated flush count per occupant for Building C, and greatest uncertainty for Building A. The significant variability between buildings in the mean daily flush count per capita broadly matches the trends observed by Melville-Shreeve et al. (2021a) and may be explained by variation in what each building is used for. Buildings that are used for teaching (A, B, D and E), for example, may have a very high capacity but a relatively low occupancy duration for each individual, and thus a low mean number of flushes per occupant; conversely, each user of a building that is predominantly used for offices (C) may spend a longer period of time in the building and therefore contribute more flushes. However, any changes in building use as a result of the pandemic (such as a building with teaching space being used only for research) may contribute further uncertainty in the daily per capita flushes and associated occupancy estimates.

**Table 2.**
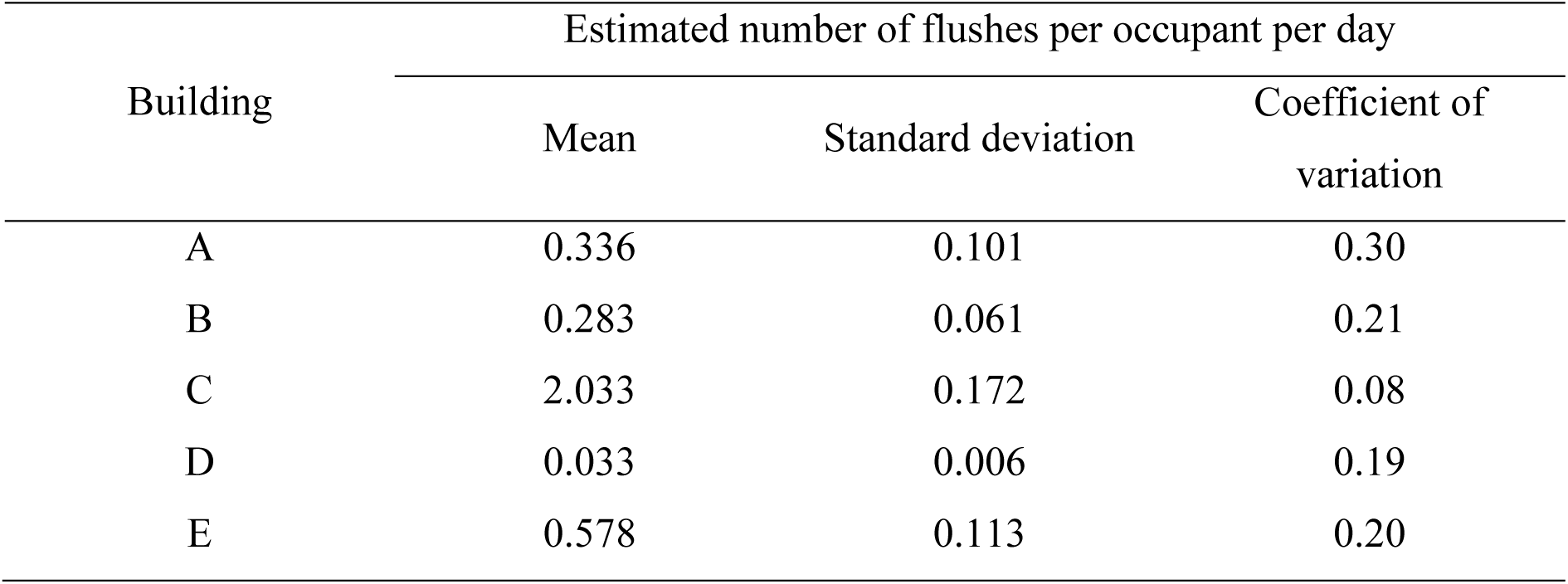
Daily per capita flush counts used to estimate dynamic population, calculated based on building capacities and flush counts during a full occupancy period

| Building | Estimated number of flushes per occupant per day |  |  |
| --- | --- | --- | --- |
|  | Mean | Standard deviation | Coefficient of variation |
| A | 0.336 | 0.101 | 0.30 |
| B | 0.283 | 0.061 | 0.21 |
| C | 2.033 | 0.172 | 0.08 |
| D | 0.033 | 0.006 | 0.19 |
| E | 0.578 | 0.113 | 0.20 |

Building-level occupancy estimates during the wastewater monitoring period, based on total flush counts (available in the Supplementary Information, Figure S4) and the estimated number of flushes per occupant per day (Table 2) are provided in Figure 7. The estimated number of occupants in each building (Figure 7a) exhibits a clear weekly pattern, with near zero occupancy in all buildings at weekends. Similarly to the campus-level population trends indicated by the wastewater biomarker loads (Figure 4a), building occupancy drops throughout December, and is lowest between the start of the Christmas closure period (CP-S) and the following start of term (T-S).

**Figure 7.**
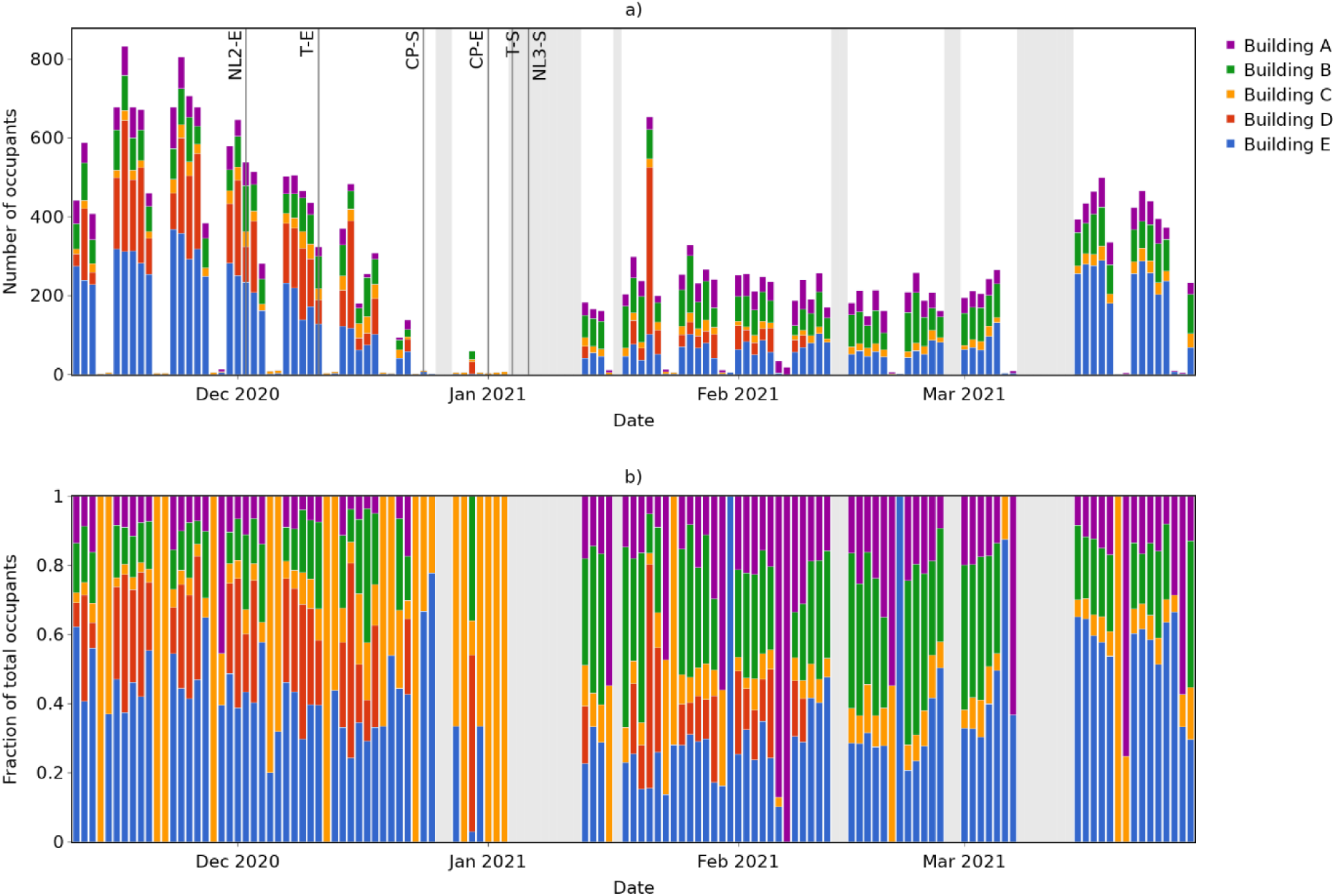
Building occupancy estimates based on flush count data: a) Absolute occupancy; and b) Relative occupancy (fraction of total occupants in monitored buildings).

Figure 7b shows how occupants are distributed between the monitored buildings and, therefore, the relative contribution of each building to the wastewater monitored for SARS-CoV-2. This is particularly insightful on days with low total occupancy numbers, showing for example that there are several days where occupants are detected only in Building C. Whilst this does not guarantee that there was no-one in the other monitored buildings, it does mean that nobody in them contributed to the wastewater being sampled, and thus these buildings can be eliminated when searching for the source of any SARS-CoV-2 detected in the wastewater on these days.

To illustrate the potential benefit of these building-level occupancy estimates for SARS-CoV-2 source identification, Figure 8 shows the wastewater SARS-CoV-2 metrics for the one-month period in which detection was most frequent (and, thus, source identification is of greatest potential benefit) overlaid on corresponding occupancy estimates.

**Figure 8.**
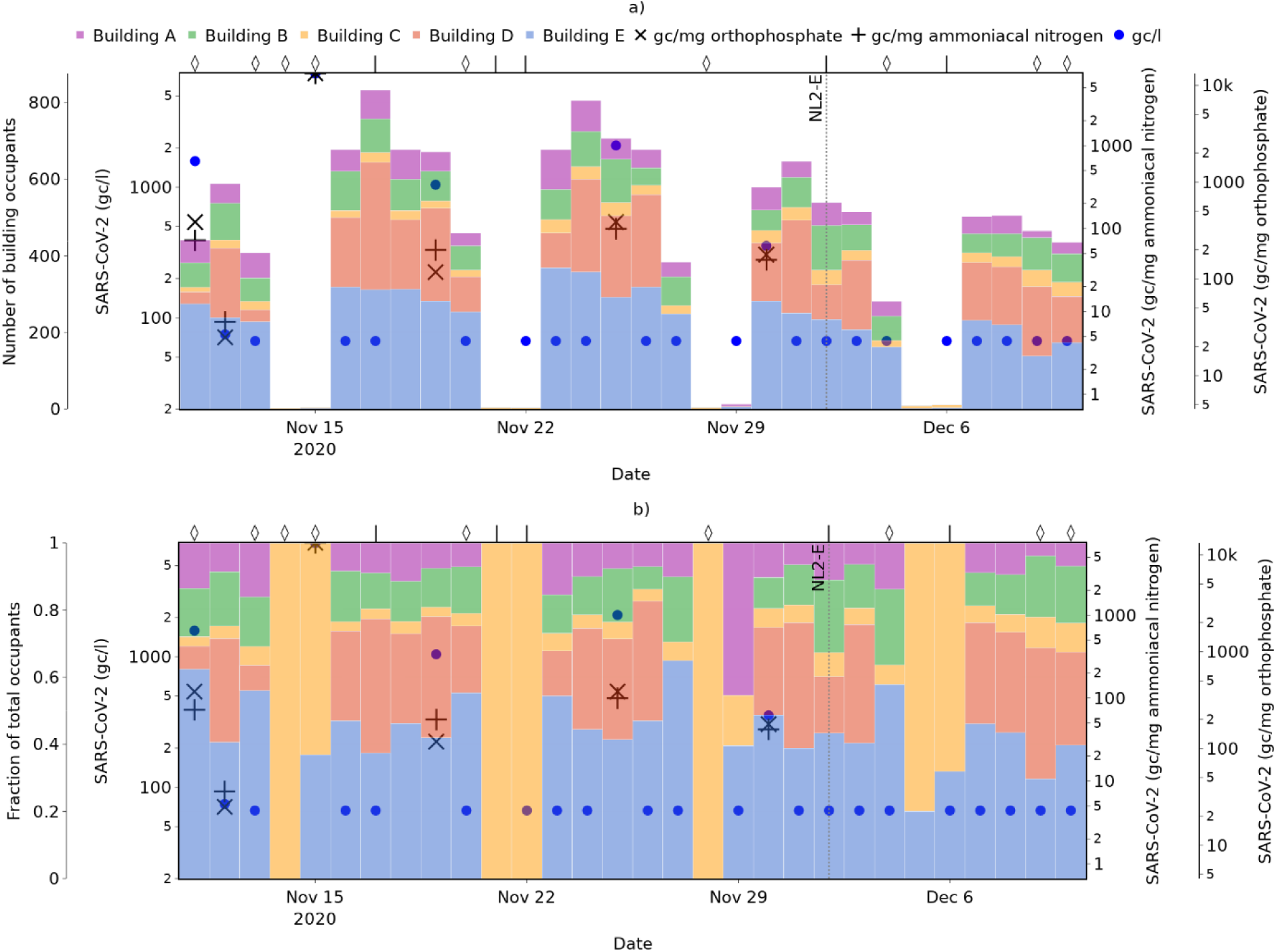
Estimated occupancy of buildings with monitored flush counts and measures of site-level SARS-CoV-2 in wastewater during the period 11^th^ November to 11^th^ December 2020, showing a) absolute occupancy; and b) relative occupancy. Wet weather days are indicated with a ‘⋄’ symbol, and dry weather days with a ‘|’.

On 15^th^ November 2020, the highest wastewater SARS-CoV-2 levels seen at the university were recorded. Figure 8b shows that buildings A, B, and D can be removed as candidates for the source, since their estimated occupancy on this day was zero. Whilst there is uncertainty in most occupancy estimates due to variation in the building-specific daily flushes per capita values, there is greater certainty of any occupant that may have been present not contributing to the wastewater on days where the estimated occupancy is zero (i.e. zero flushes). However, it is noted that flush data was not captured at wheelchair-accessible washrooms, and thus the possibility of users of these buildings contributing to the wastewater cannot be absolutely ruled out.

Furthermore, Figure 8a shows that the occupancy of all buildings with monitored flush counts were very low on this day. If it is assumed that the occupancy of these buildings is broadly representative of occupancy across the campus on this day, then this suggests that the actual number of people infected may also be very low, despite both the SARS-CoV-2 concentration and the population normalised values being very high (since the smaller the population, the higher the per capita value resulting from a given SARS-CoV-2 load).

On days where specific buildings cannot be eliminated based on a zero-occupancy estimate, building-level occupancy information may still aid efforts to trace the source of SARS-CoV-2 detected by enabling identification of buildings with the greatest occupancy and greatest wastewater contribution. Figure 8b shows that on 11^th^ November, for example, over 60% of the total occupants of five monitored buildings were in Building E – hence, there is greater probability of locating the infected individual(s) in this building, and this should be a higher priority for targeted testing if capacity is limited.

### 3.4. Future opportunities

Multiple opportunities are identified to add value to outputs set out in this study. Specifically, the installation of flush monitoring technology across all washrooms upstream of the autosampler could enable a more definitive set of conclusions to be drawn. Disaggregation of flush counts from male and female washrooms may also improve accuracy when assessing the relative contribution of occupants in different buildings to the wastewater sampled, due to the presence of (unmonitored) urinals in the male washrooms.

In addition, from an operational perspective, additional autosamplers could be installed (but remain largely offline) at the outlet from each building. These could be sampled the day after a positive signal is observed in at the main campus, enabling a single building to be pin-pointed. Such measures could in turn enable patchwork closure of buildings when prevalence exceeds a pre-defined threshold, minimising disruption in future waves of a pandemic.

## 4. CONCLUSIONS

This study has investigated the use of ammoniacal nitrogen and orthophosphate for normalisation of SARS-CoV-2 detected in wastewater, to account for the impact of highly variable populations at a near-to-source monitoring site; evaluated the impact of population normalisation on the understanding of SARS-CoV-2 prevalence trends provided by the wastewater data; demonstrated how complementary (non-wastewater) data sources can help to inform a better targeted response; and explored the potential impact of wet weather periods on the results. Key findings include:

- Population normalisation alters the trends in SARS-CoV-2 prevalence indicated by WBE and, in a near-to-source site with a highly variable population such as a university campus, it can reveal significant differences in prevalence between days where recorded SARS-CoV-2 concentrations are very similar. Population normalisation, therefore, is considered critical for providing a comprehensive understanding of the results from WBE when population size is highly variable.
- Normalisation using either ammoniacal nitrogen or orthophosphate is similarly beneficial, with both providing a similar (but not identical) understanding of population dynamics and trends in population normalised SARS-CoV-2 in the wastewater. This indicates that multiple biomarkers that are of questionable reliability for population normalisation at a STW level due to their presence in industrial discharges can be appropriate for near-to-source studies.
- Agreement between population estimates based on different biomarkers is greatest when wet weather days are omitted, indicating that confidence in the results of population normalisation should be greatest when the weather is dry. However, as there is still a reasonable level of agreement on wet days, these can still provide valuable information.
- Use of flush count data to estimate the occupancy of different buildings in within the near-to-source site can enable priority locations for targeted testing to be identified when SARS-CoV-2 is detected in the wastewater. This is particularly beneficial on low occupancy days when no flushes are recorded in some buildings, so it is known with certainty that no occupants of these buildings contributed to the wastewater in which SARS-CoV-2 was detected.
- Technically feasible strategies to further advance this study have been set out. Such solutions focus on yet more granular data acquisition including a wider deployment of flush monitoring and short term autosampling being added at a building-level when the main campus data suggests increased prevalence.

Lastly, it is noted that there were restrictions in place on mobility and/or student activities, along with guidance to ‘work from home where possible’, for the majority this study period, due to the COVID-19 pandemic. As such, the number of people using the main campus site (and the difference between high and low occupancy periods) was considerably lower than usual. For near-to-source sites with higher variability in population – and for the case study site as restrictions are lifted and the number of people using the campus increases – the importance of population normalisation is expected to be even greater.

## Data Availability

Raw data are currently unavailable for distribution; all data generated are included in the manuscript or supplementary information.

## ACKNOWLEDGEMENTS

This paper uses Environment Agency rainfall data licensed under the Open Government Licence v3.0. The sampling, testing and data analysis of wastewater in England is funded by the United Kingdom Government (Department of Health and Social Care).

## DISCLAIMER

The views expressed in this paper are those of the authors and do not necessarily reflect the views or policies of the Department of Health and Social Care.

