## Supplementary Information for "Building knowledge of university campus population dynamics to enhance near-to-source sewage surveillance for SARS-CoV-2 detection"

#### RELATIONSHIP BETWEEN SARS-COV-2 CONCENTRATION AND DAILY LOAD PER CAPITA FOR DIFFERENT POPULATIONS AND BASE FLOWS

The theoretical relationship between SARS-CoV-2 concentration ($C$, gc/l) and the daily SARS-CoV-2 per capita ($L$, gc/d/capita) is expressed as a function of the population ($N$), daily wastewater production per capita ($q$, l/d/capita) and the base flow in accordance ($Q_B$, l/d) with Eq. S1. The daily wastewater production per capita is assumed to be constant and the base flow consists of all wastewater that is not accounted for in the per capita production value (e.g. due to runoff, ingress and industrial discharges). *LN* is the total SARS-CoV-2 load per day (gc/d) and ($Nq + Q_B$) the total wastewater flow rate.

*Eq. S1*

$$C = \frac{LN}{Nq + Q_B}$$

### RELATIVE POPULATION ESTIMATES BASED ON METERED WATER AND ELECTRICITY SUPPLY

Relative population is estimated using a) water supply data and b) electricity supply data, based on the assumption that these values are linearly related to population and daily per capita usage is constant. Daily metered supply ($M_d$) is assumed to exhibit the following relationship with population:

*Eq. S2*

$$M_d = N_d m + m_B$$

Where $N_d$ is the population on day *d*, *m* is the metered supply per capita, and $m_B$ is the base supply (independent of population).

The base supply is estimated using daily metered supply values from August 2020, based on the assumption that population is zero during this period. The metered supply per capita is then estimated using daily metered supply values from the second lockdown, based on the assumption that population is static during this period, and an assumed population of 1. This then enables estimation of dynamic population relative to the lockdown period (rather than absolute population). Example results for Building E are shown in Figure S1 and Figure S2, illustrating the poor agreement between the estimates and high uncertainty.

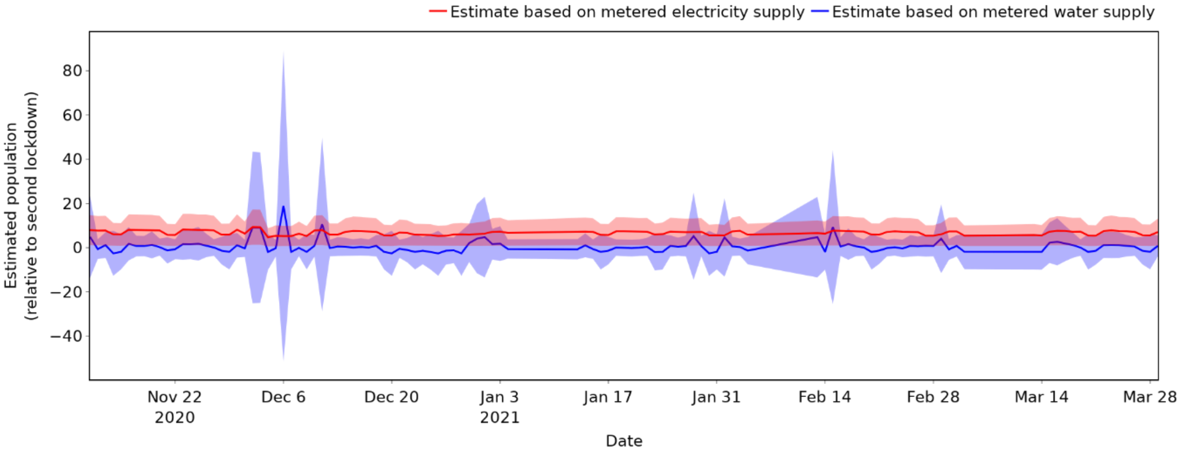


*Figure S1. Relative population estimates (mean and standard deviation) for Building E, based on metered water and electricity supply.*

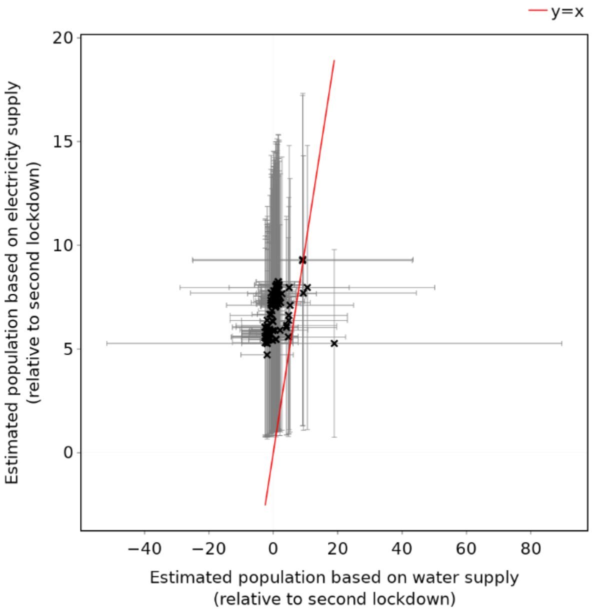


*Figure S2. Relationship between relative population estimates for Building E based on metered water and electricity supply. Error bars indicate standard deviation.*

### DATA AVAILABLITY AND COMPLETENESS

Flush data completeness at a washroom level (prior to omission of washrooms and/or days with insufficient completeness) is shown in Figure S3**Error! Reference source not found.**.

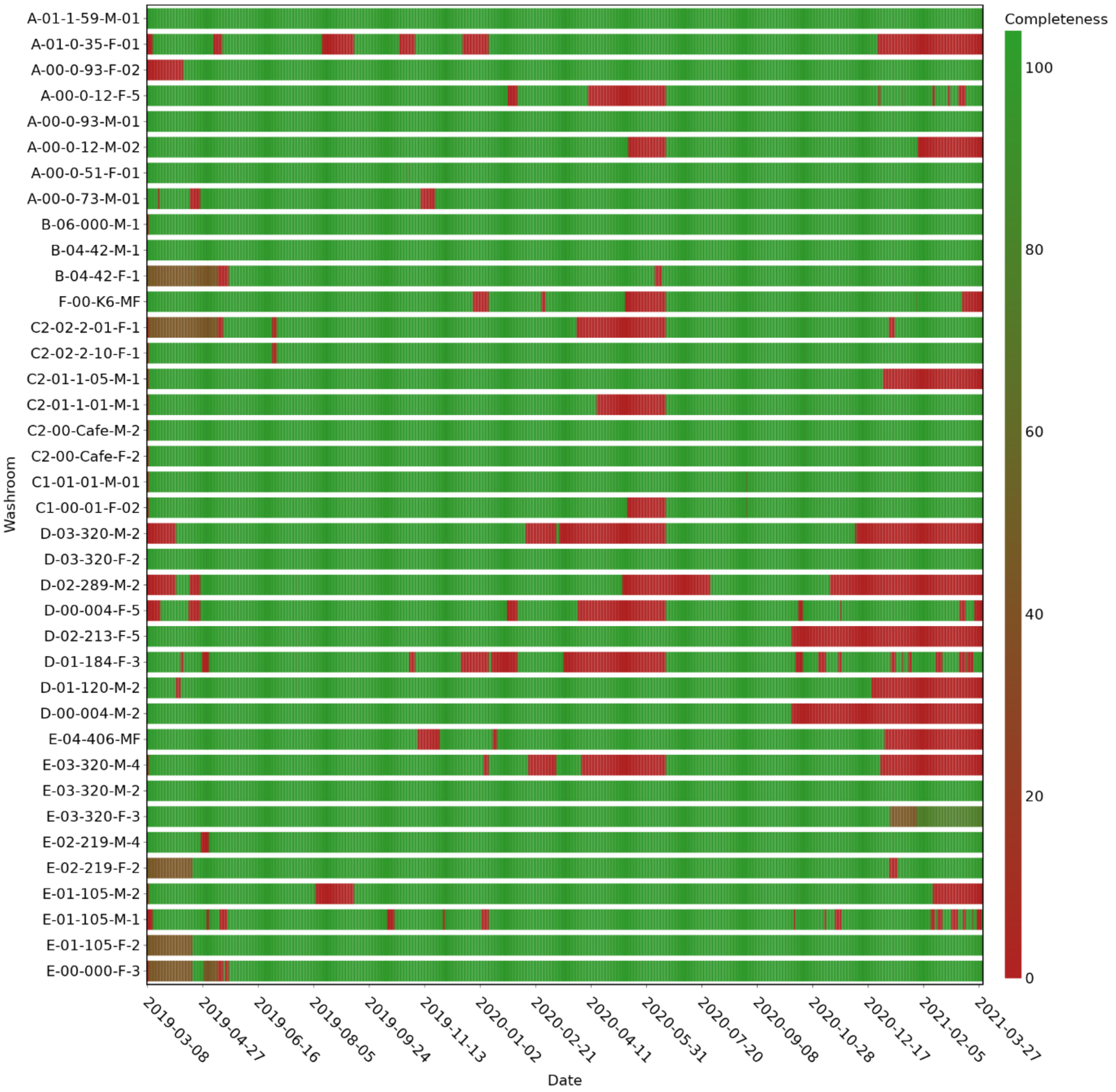


*Figure S3. Raw flush data completeness at a washroom level*

### FLUSH COUNTS ADJUSTED FOR CLEANING

Total flush counts for each building during the wastewater monitoring period, before and after removal of flushes attributed to cleaning, are shown in Figure S4. The percentage of the total flush count attributed to cleaning in each building is summarised in Table S1. The total flush count across all buildings, after adjustment for cleaning, is shown in Figure S5 and Figure S6.

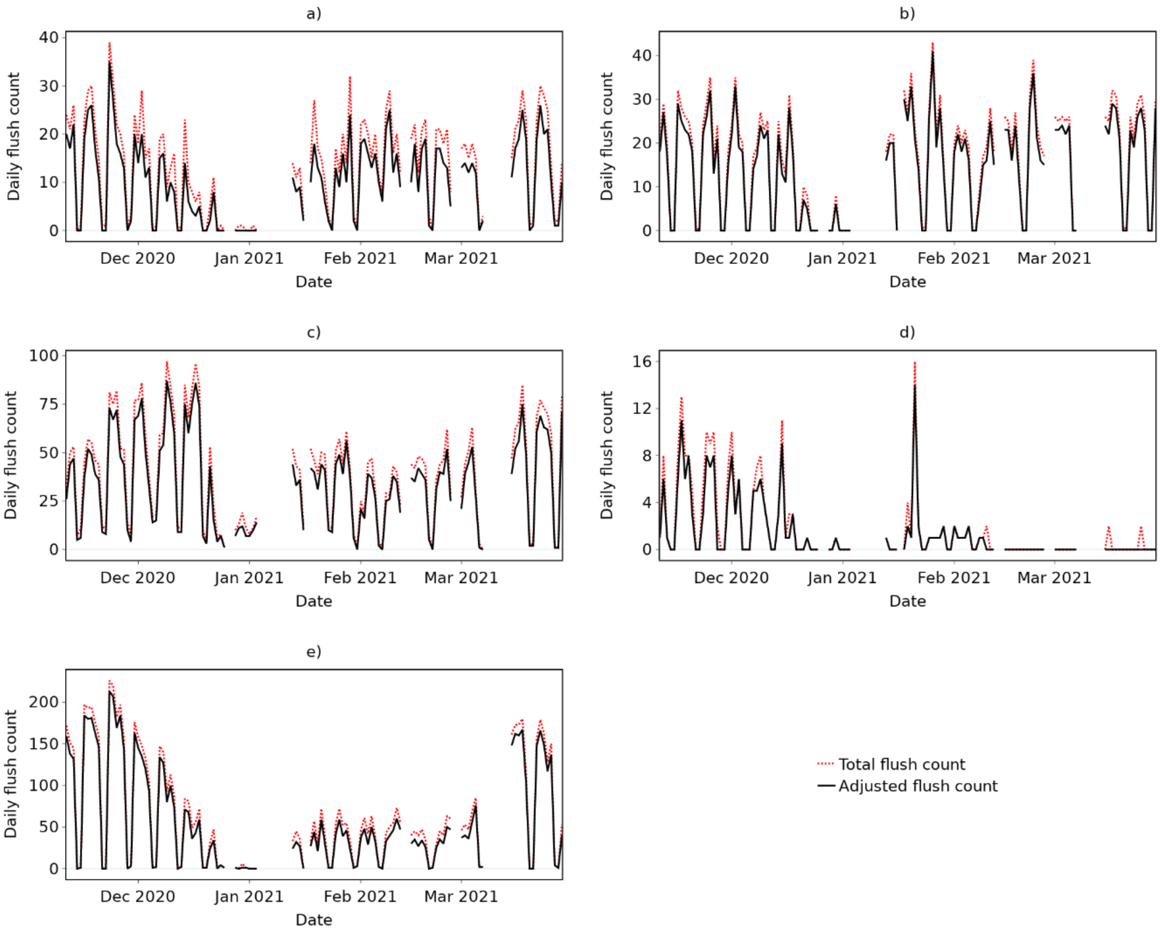


*Figure S4. Total and adjusted flush counts for buildings a) A; b) B; c) C; d) D; and e) E.*

*Table S1. Percentage of total flush count during wastewater monitoring period attributed to cleaning in each building.*

| Building | Flushes attributed to cleaning |
|---|---|
| A | 23.1% |
| B | 10.7% |
| C | 14.0% |
| D | 20.1% |
| E | 12.1% |

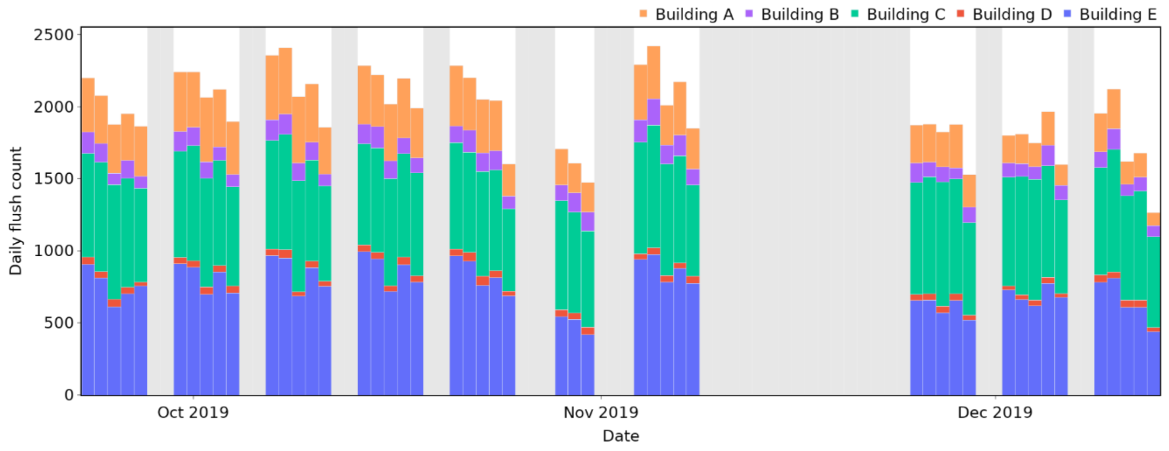


*Figure S5. Total adjusted flush counts across monitored buildings during assumed full occupancy period*

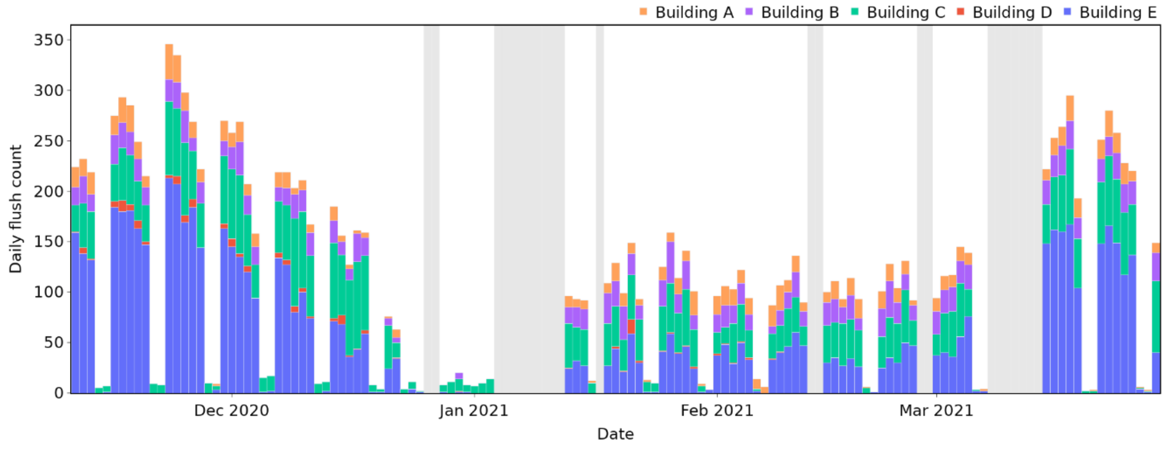


*Figure S6. Total adjusted flush counts across monitored buildings during the wastewater monitoring period*

### CORRELATIONS BETWEEN WASTEWATER METRICS

*Table S2. Summary of statistics related to the correlation between wastewater metrics. Confidence intervals are calculated using a significance level of 0.05.*

| | Parameter 1 (log10) | Parameter 2 (log10) | Number of samples | Pearson correlation coefficient | 2-tailed p-value | Confidence interval Lower | Confidence interval Upper |
|---|---|---|---|---|---|---|---|
| All samples | mg ammoniacal nitrogen / day | mg orthophosphate / day | 95 | 0.792 | 0.0000 | 0.702 | 0.857 |
| | SARS-CoV-2 gc/l | SARS-CoV-2 gc/mg ammoniacal nitrogen | 12 | 0.899 | 0.0001 | 0.672 | 0.972 |
| | SARS-CoV-2 gc/l | SARS-CoV-2 gc/mg orthophosphate | 11 | 0.914 | 0.0001 | 0.696 | 0.978 |
| | SARS-CoV-2 gc/mg ammoniacal nitrogen | SARS-CoV-2 gc/mg orthophosphate | 11 | 0.981 | 0.0000 | 0.927 | 0.995 |
| Dry weather samples only | mg ammoniacal nitrogen / day | mg orthophosphate / day | 27 | 0.917 | 0.0000 | 0.824 | 0.962 |
| | SARS-CoV-2 gc/l | SARS-CoV-2 gc/mg ammoniacal nitrogen | 4 | 0.930 | 0.0696 | -0.290 | 0.999 |
| | SARS-CoV-2 gc/l | SARS-CoV-2 gc/mg orthophosphate | 4 | 0.952 | 0.0484 | -0.111 | 0.999 |
| | SARS-CoV-2 gc/mg ammoniacal nitrogen | SARS-CoV-2 gc/mg orthophosphate | 4 | 0.997 | 0.0028 | 0.867 | 1.000 |
